# Practical considerations for measuring the effective reproductive number, *R_t_*

**DOI:** 10.1101/2020.06.18.20134858

**Authors:** Katelyn M. Gostic, Lauren McGough, Edward B. Baskerville, Sam Abbott, Keya Joshi, Christine Tedijanto, Rebecca Kahn, Rene Niehus, James Hay, Pablo M. De Salazar, Joel Hellewell, Sophie Meakin, James Munday, Nikos I. Bosse, Katharine Sherrat, Robin N. Thompson, Laura F. White, Jana S. Huisman, Jérémie Scire, Sebastian Bonhoeffer, Tanja Stadler, Jacco Wallinga, Sebastian Funk, Marc Lipsitch, Sarah Cobey

## Abstract

Estimation of the effective reproductive number, *R_t_*, is important for detecting changes in disease transmission over time. During the COVID-19 pandemic, policymakers and public health officials are using *R_t_* to assess the effectiveness of interventions and to inform policy. However, estimation of *R_t_* from available data presents several challenges, with critical implications for the interpretation of the course of the pandemic. The purpose of this document is to summarize these challenges, illustrate them with examples from synthetic data, and, where possible, make recommendations. For near real-time estimation of *R_t_*, we recommend the approach of Cori et al. (2013), which uses data from before time t and empirical estimates of the distribution of time between infections. Methods that require data from after time *t*, such as Wallinga and Teunis (2004), are conceptually and methodologically less suited for near real-time estimation, but may be appropriate for retrospective analyses of how individuals infected at different time points contributed to spread. We advise against using methods derived from Bettencourt and Ribeiro (2008), as the resulting *R_t_* estimates may be biased if the underlying structural assumptions are not met. Two key challenges common to all approaches are accurate specification of the generation interval and reconstruction of the time series of new infections from observations occurring long after the moment of transmission. Naive approaches for dealing with observation delays, such as subtracting delays sampled from a distribution, can introduce bias. We provide suggestions for how to mitigate this and other technical challenges and highlight open problems in *R_t_* estimation.

**Author summary:** The effective reproductive number, *R_t_*, is a key epidemic parameter used to assess whether an epidemic is growing, shrinking or holding steady. *R_t_* estimates can be used as a near real-time indicator of epidemic growth or to assess the effectiveness of interventions. But due to delays between infection and case observation, estimating *R_t_* in near real-time, and correctly inferring the timing of changes in *R_t_* is challenging. Here, we provide an overview of challenges and best practices for accurate, timely *R_t_* estimation.

## Introduction

The effective reproductive number, denoted *R_e_* or *R_t_*, is the expected number of new infections caused by an infectious individual in a population where some individuals may no longer be susceptible. Estimates of *R_t_* are used to assess how changes in policy, population immunity, and other factors have affected transmission at specific points in time [1-5]. The effective reproductive number can also be used to monitor near real-time changes in transmission [6-10]. For both purposes, estimates need to be accurate and correctly represent uncertainty, and for near real-time monitoring, they also need to be timely.

We consider two potential forms of bias in *R_t_* estimates, systematic over- or underestimation and temporal inaccuracy. Misspecification of the generation interval is a large potential source of over- or underestimation, and we find that *R_t_* estimates are most prone to this kind of bias when the true value is substantially greater or less than one. This situation might arise at the beginning of the COVID-19 pandemic (when *R_t_* is relatively high) or after particularly effective interventions (when it might be low). Over or underestimation would have particularly strong practical consequences near the control threshold of *R_t_* = 1, but the biases we observe are smallest in absolute terms in this range.

Another challenge is that depending on the methods used, *R_t_* estimates may be leading or lagging indicators of the true value [4, 11], even measuring transmission events that occurred days or weeks ago if the data are not properly adjusted. Temporal inaccuracy in *R_t_* estimation is particularly concerning when trying to infer how changes in behavior have affected transmission [1-5]. Temporal inaccuracy, for instance in the estimation of the date on which *R_t_* falls below one, is a focus of this Perspective. We find that it has several possible causes and can be difficult to avoid.

This perspective focuses on the three main empirical methods to estimate *R_t_* [12-15]. As an alternative to the methods reviewed here, it is possible to infer changes in transmission using a dynamical compartment model (e.g. [3, 16-18]). The accuracy and timeliness of *R_t_* estimates obtained in this way should be assessed on a case-by-case basis, given sensitivity to model structure and data availability.

We use synthetic data to compare the accuracy of three common empirical methods to estimate *R_t_*, first under ideal conditions, in the absence of parametric uncertainty and with all infections observed at the moment they occur. This idealized analysis is intended to illustrate the inputs needed to estimate *R_t_* accurately, to highlight the intrinsic differences between the methods, and to examine specific causes of bias and temporal inaccuracy one by one. However, we emphasize that our idealized analyses overestimate the potential accuracy of *R_t_* estimates obtained from real-world data, even if best practice are followed. The results show that the method of Cori et al. [14] is best for near real-time estimation of *R_t_*. For retrospective analysis, the methods of Cori et al. or of Wallinga and Teunis may be appropriate, depending on the aims.

Later we add realism and address practical considerations for working with imperfect data. These analyses emphasize potential errors introduced by uncertainty in the intrinsic generation interval and imperfect case observation, and also the need to adjust for delays in case observation, right truncation, and the need to choose an appropriate smoothing window given the sample size. Finally, we emphasize that most off-the-shelf tools leave it up to the user to account for these five sources of uncertainty when calculating confidence intervals. Failure to propagate uncertainty in *R_t_* estimates can lead to over-interpretation of the results and could falsely imply that confidence or credible intervals have crossed the critical threshold.

## Synthetic data

We used synthetic data to compare three common *R_t_* estimation methods. Synthetic data were generated from a deterministic or stochastic SEIR model in which the transmission rate drops and spikes abruptly, representing the adoption and lifting of public health interventions. Results were similar whether data were generated using a deterministic or stochastic model. For simplicity, we show deterministic outputs throughout the document, except in the section on smoothing windows, where stochasticity is a conceptual focus.

In our model, all infections are locally transmitted, but all three of the methods we test can incorporate cases arising from importations or zoonotic spillover [12, 13, 15]. Estimates of *R_t_* are likely to be inaccurate if a large proportion of cases involve transmission outside the population. This situation could arise when transmission is low (e.g., at the beginning or end of an epidemic) or when *R_t_* is defined for a population that is connected to others via migration.

A synthetic time series of new infections (observed at the *S → E* transition) was input into the *R_t_* estimation methods of Wallinga and Teunis, Cori et al., and Bettencourt and Ribeiro [12-14]. Following the published methods, we also tested the Wallinga and Teunis estimator using a synthetic time series of symptom onset events, extracted daily from the *E → I* transition. In the synthetic data, the generation interval followed a gamma distribution with shape 2 and rate 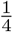, which is the sum of exponentially distributed residence times in compartments *E* and *I*, each with mean of 4 days [19]. The methods of Cori et al. and of Wallinga and Teunis can accommodate any positive, discrete generation interval distribution [12, 14, 20]. We chose a discretized, gamma-distributed generation interval for our simulations, which is similar in shape to the interval of COVID-19 [21, 22]. However, the method of Bettencourt and Ribeiro implicitly assumes the generation interval follows an exponential distribution, as in an SIR model with no latent period [13, 19], and thus could not match the assumptions of the synthetic data.

In the synthetic data, *R_0_* was set to 2.0 initially, then to 0.8 and 1.15, to simulate the adoption and later the partial lifting of public health interventions. To mimic estimation in real-time, we truncated the time series at *t* = 150, before the end of the epidemic. Estimates from the methods of Wallinga and Teunis and Cori et al. were obtained using the R package EpiEstim [20]. Estimates based on the method of Bettencourt and Ribeiro were obtained by translating code from [6, 23] to the Stan language [24]. We initially assumed all infections were observed. Unless otherwise noted, the smoothing window was set to 1 day (effectively, estimates were not smoothed). To mimic the timescale of observations, we used daily time steps when generating synthetic data and performing analyses.

### Comparison of common methods

The effective reproductive number at time t can be defined in two ways: as the instantaneous reproductive number or as the case reproductive number [14, 25]. The instantaneous reproductive number measures transmission at a specific point in time, whereas the case reproductive number measures transmission by a specific cohort of individuals (Fig. 1). (A cohort is a group of individuals with the same date of infection or the same date of symptom onset.) The case reproductive number is useful for retrospective analyses of how individuals infected at different time points contributed to spread. It is a more natural choice for analyses that consider heterogeneity among individuals. For example, the case reproductive number of Wallinga and Teunis can be adapted to incorporate data on observed transmission chains [12, 21, 26] or to produce age-structured *R_t_* estimates, given an age-structured contact matrix [27]. The instantaneous reproductive number is more appropriate for analyses estimating the reproductive number of the infected population on specific dates, especially when aiming to study how interventions or other extrinsic factors have affected transmission at a given point in time.

**Fig 1.**
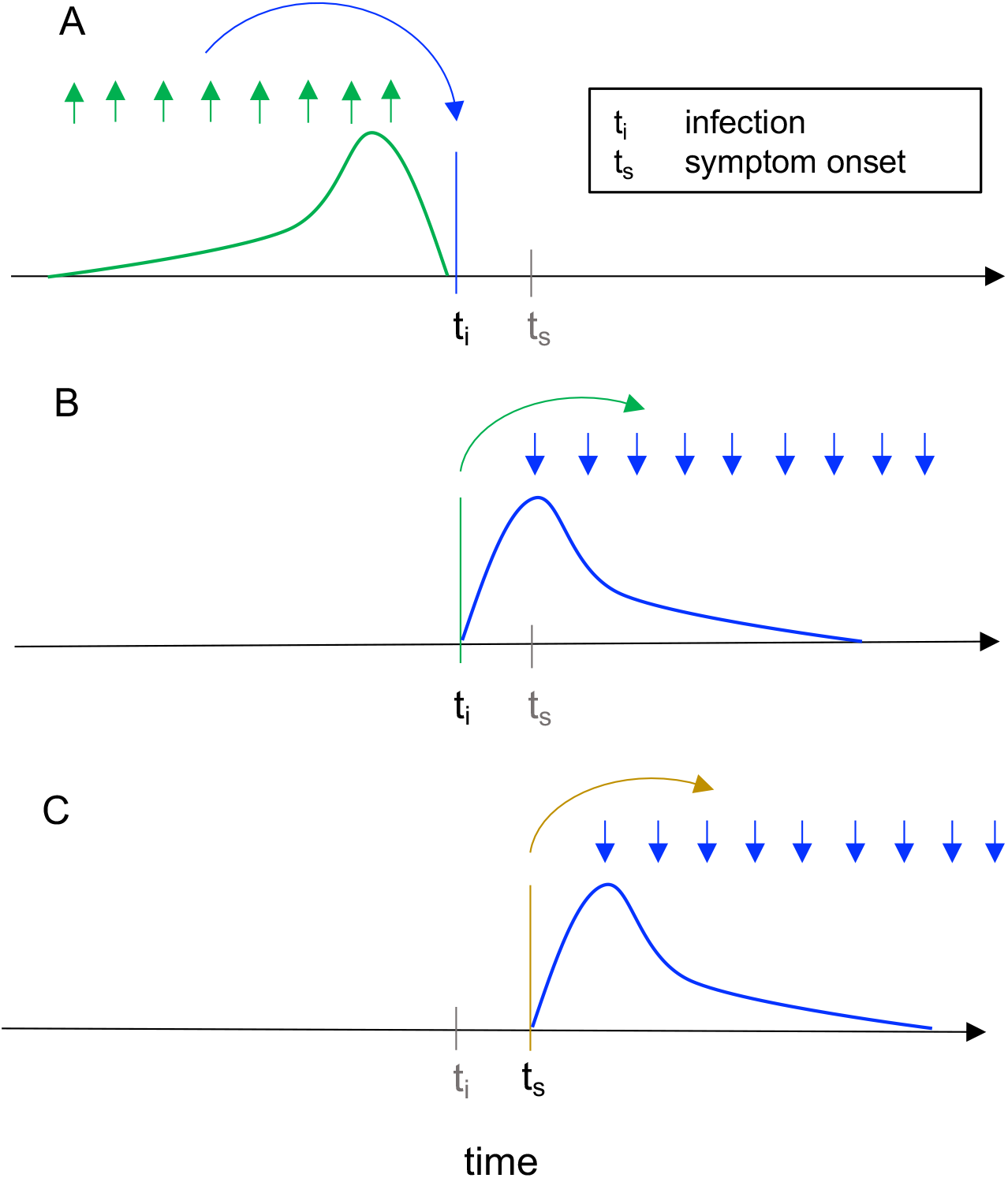
Instantaneous reproductive number as estimated by the method of Cori et al. versus cohort reproductive number estimated by Wallinga and Teunis. For each definition of *R_t_*, arrows show the times at which infectors (upwards), and their infectees (downwards) appear in the data. Curves show the generation interval distribution (A, B), or serial interval distribution (C). A: The instantaneous reproductive number quantifies the number of new infections incident at a single point in time (*t_i_* blue arrow), relative to the number of infections incident in the previous generation (green arrows), and their current infectiousness (green curve). This figure illustrates the method of Cori et al. B & C: The case reproductive number of Wallinga and Teunis is the average number of new infections that an individual who becomes (B) infected on day *t_i_* (green arrow) or (C) symptomatic on day *t_s_* (yellow arrow) will eventually go on to cause (blue downward arrows show timing of daughter cases). The first definition applies when estimating the case reproductive number using inferred times of infection, and the second applies when using data on times of symptom onset.

More formally, the instantaneous reproductive number is defined as the expected number of secondary infections occurring at time *t*, divided by the number of infected individuals, each scaled by their relative infectiousness at time *t* (an individual’s relative infectiousness is based on the generation interval, and time since infection) [14, 25]. The instantaneous reproductive number can be calculated exactly for a compartment model (SIR or SEIR) as follows, where *β*(*t*) is the time-varying transmission rate, *S*(*t*) the fraction of the population that is susceptible, and D the mean duration of infectiousness:

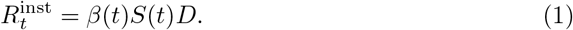

The methods of Cori et al. [14, 15] and methods adapted from Bettencourt and Ribeiro [6, 13, 23] estimate the instantaneous reproductive number from observations. We tested their accuracy under idealized conditions, assuming perfect knowledge of the generation interval and delay distributions used to generate the synthetic data (Fig. 2 and S2 Fig).

**Fig 2.**
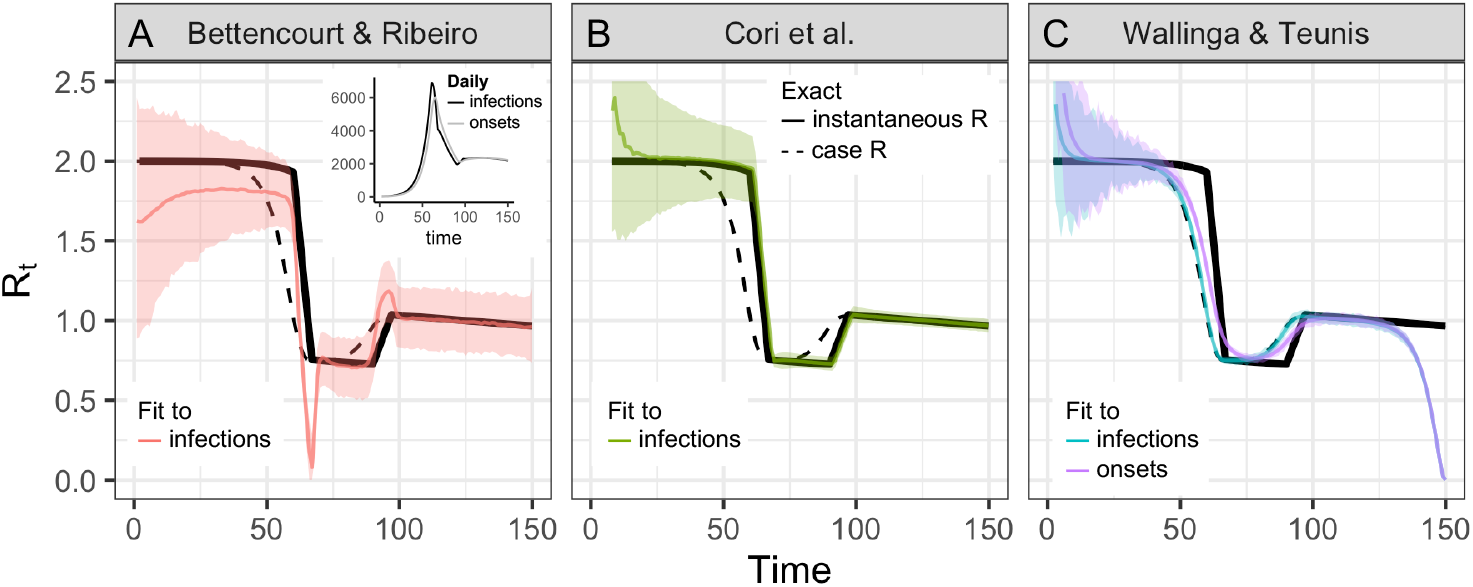
Accuracy of *R_t_* estimation methods given ideal, synthetic data. Solid and dashed black line shows the instantaneous and case reproductive numbers, respectively, calculated from synthetic data. Colored lines show estimates and confidence or credible intervals. To mimic an epidemic progressing in real-time, the time series of infections or symptom onset events up to *t* = 150 was input into each estimation method (inset). Terminating the time series while *R_t_* is falling or rising produces similar results S2 Fig. **(A)** By assuming a SIR model (rather than SEIR, the source of the synthetic data), the method of Bettencourt and Ribeiro systematically underestimates *R_t_* when the true value is substantially higher than one. The method is also biased as transmission shifts. **(B)** The Cori method accurately measures the instantaneous reproductive number. **(C)** The Wallinga and Teunis method estimates the cohort reproductive number, which incorporates future changes in transmission. Thus, the method produces *R_t_* estimates that lead the instantaneous effective reproductive number and becomes unreliable for real-time estimation at the end of the observed time series without adjustment for right truncation [4, 30]. In (A, B) the colored line shows the posterior mean and the shaded region the 95% credible interval. In (C) the colored line shows the maximum likelihood estimate and the shaded region the 95% confidence interval.

The method of Cori et al. estimates *R_t_* as

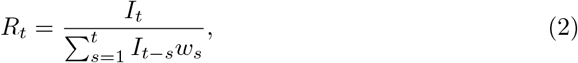

where *I_t_* is the number of infections incident on day t and *w_s_* is generation interval, or the probability that s days separate the moment of infection in an index case and a daughter case [14]. Conceptually, this estimator describes the number of new infections incident on day t relative to the number (*I_t−s_*) and current infectiousness (*w_s_*) of individuals who became infected s days in the past, and who many now be shedding virus.

The only parametric assumption required by this method is the form of the generation interval. The standard assumption is that *w_s_* follows a discretized gamma distribution [14, 20], but the estimator accepts any parametric or empirical discrete distribution with support on positive values (the same is true of the Wallinga and Teunis method [12]). Thus, when testing the method on COVID-19-like epidemic processes, we could specify the gamma-distributed generation interval of the synthetic data perfectly. When tested under idealized conditions, we found that the method of Cori et al. accurately estimated *R_t_*, even tracking abrupt changes (Fig. 2). This is the method we recommend for estimation of the instantaneous reproductive number.

Bettencourt and Ribeiro [13] derive an approximate relationship between the *R_t_* and the exponential growth rate of the epidemic, where *g* is the mean generation time:

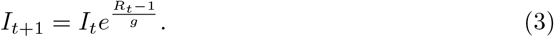

Under the assumption that *R_t_* evolves through time as a Gaussian process, equation 3 facilitates efficient Bayesian *R_t_* estimation [23]. The key disadvantage of this method is that equation 3 is derived from an SIR model, and therefore implicitly assumes the generation interval follows an exponential distribution [19], whereas empirically, generation intervals can be heavy or light tailed, including for COVID-19 [21, 22, 28]. Because equation 3 underestimates variability in the gamma-distributed generation interval of our SEIR-type synthetic data, we find that this method produces biased *R_t_* estimates, especially when *R_t_* is substantially higher than one (Fig. 2A). These biases are consistent with established theory, in which the coefficient of variation of the generation interval modulates the relationship between exponential growth rate and the reproductive number of an epidemic [19].

In its current form, we do not recommend using the method of Bettencourt and Ribeiro given that unrealistic structural assumptions lead to bias. However, a generalized version capable of accommodating more realistic generation intervals, which implicitly involves different assumptions about the underlying epidemic process [19, 29], could provide several advantages. When implemented as a Gaussian process [23], we found that the Bettencourt and Ribeiro method was computationally efficient. Also, because it penalizes large jumps in *R_t_* across consecutive time steps, it returns smoother estimates than the method of Cori et al., which is advantageous if unmodeled reporting effects, rather than bursts in transmission, are the dominant cause of variability in daily observations.

Finally, the **case or cohort reproductive number** is the expected number of secondary infections that an individual who becomes infected at time t will eventually cause as they progress through their infection [14, 19, 25] (Fig. 1B, C). The case reproductive number 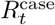 can be calculated exactly at time t within the synthetic data as the convolution of the generation interval distribution *w*(·) and the instantaneous reproductive number, 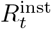, described in Equation 1 [19],

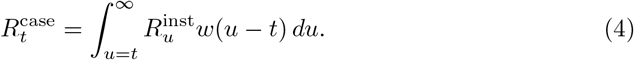

The method of Wallinga and Teunis [12] estimates the case reproductive number from observations. The first step is to estimate the likelihood that case *j* (infected at time *t_j_*) infected case *i*, relative to the likelihood that any other case in the data infected case *i*:

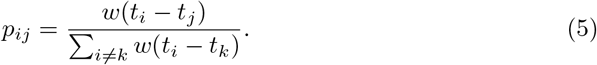

Then, the individual reproductive number of case *j* is defined as

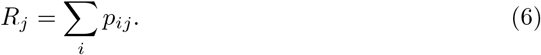

The case reproductive number at time t is defined as the expected value of *R_j_* for all individuals infected at time *t*. Like the method of Cori et al., the only parametric assumption required by the method of Wallinga and Teunis is the discrete distribution of the generation interval. An assumption common to the three tested methods is that all infections are observed. Below we discuss the consequences of partial observation, including that confidence or credible intervals around *R_t_* estimates do not account for uncertainty from partial observation.

Practically speaking, there are several important differences between the case reproductive number (estimated by Wallinga and Teunis) and the instantaneous reproductive number (estimated by Cori et al. or Bettencourt and Ribeiro). First, estimators of the instantaneous reproductive number were partly developed for near real-time estimation and only use data from before time t (Fig. 1A). Under ideal conditions without observation delays and a window size of one day, neither method is affected by the termination of the synthetic time series at t = 150 (Figs. 2 A&B). These methods are similarly robust if the time series ends while *R_t_* is rapidly falling (S2 FigA) or rising (S2 FigB). (Below we discuss more realistic conditions, e.g., in which data at the end of a right-truncated time series are incomplete due to observation delays.)

Unlike the instantaneous reproductive number, the case reproductive number is inherently forward-looking (Fig. 1B, C): near the end of a right-truncated time series, it relies on data that have not yet been observed. Extensions of the Wallinga and Teunis method can be used to adjust for these missing data and to obtain accurate *R_t_* estimates to the end of a truncated time series [4, 30]. But as shown in Fig. 2C, without these adjustments the method will always underestimate *R_t_* at the end of the time series, even in the absence of reporting delays. Mathematically, this underestimation occurs because calculating the case reproductive number involves a weighted sum across transmission events observed after time t. Time points not yet observed become missing terms in the weighted sum. Similarly, any infections that occurred before the first observed date are missing terms in the denominator of the Cori et al. estimator, and so the method of Cori et al. often overestimates *R_t_* early in the time series.

Effectively, the case reproductive number is shifted forward in time relative to the instantaneous reproductive number of Cori et al. (S3 Fig). This temporal shift occurs whether or not a smoothing window is used. The case reproductive number produces leading estimates of changes in the instantaneous reproductive number (Fig. 2, S3 Fig) because it uses data from time points after t, whereas the instantaneous reproductive number uses data from time points before t (Fig. 1). Shifting the case reproductive number back in time by the mean generation interval usually provides a good approximation of the instantaneous reproductive number [2], because the case reproductive number is essentially a convolution of the instantaneous reproductive number and the generation interval (Equation 4) [19]. The case reproductive number generally changes more smoothly than the instantaneous reproductive number [25] (Fig. 2B, C), but if a smoothing window is used, the estimates become more similar in shape and smoothness. Overall, for real-time analyses aiming to quantify the reproductive number at a particular moment in time or to infer the impact of changes in policy, behavior, or other extrinsic factors on transmission, the instantaneous reproductive number will provide more temporally accurate estimates and is most appropriate. The case reproductive number of Wallinga and Teunis considers the reproductive number of specific individuals, and therefore is more appropriate for analyses aiming to incorporate individual-level covariates such as age [27] or transmission cluster membership [12] when estimating *R_t_*.

### Summary

- The Cori method most accurately estimates the instantaneous reproductive number in real-time. It uses only past data and minimal parametric assumptions.
- The method of Wallinga and Teunis estimates a slightly different quantity, the case or cohort reproductive number. The case reproductive number is conceptually less appropriate for real-time estimation but may be useful in retrospective analyses, especially those involving individual-level covariates.
- Methods adapted from Bettencourt and Ribeiro [6, 13] can lead to biased *R_t_* estimates if the underlying structural assumptions are not met.

## Generation interval misspecification

When estimating *R_t_* from observed data, misspecification of the generation interval is a large potential source of bias. Regardless of the method used, *R_t_* estimates are not only sensitive to the mean generation time but also to the variance and form of the generation interval distribution [19].

The renewal equation is a cornerstone of demographic theory and forms the mathematical backbone of the *R_t_* estimators described above [19]. Within the renewal equation, the generation interval mechanistically links the reproductive number R to observables such as the epidemic growth rate r or the number of new infections per day [19]. Wallinga and Lipsitch [19] describe how the exponential growth rate of the Bettencourt and Ribeiro estimator (their equation 3.1) and continuous-time equivalents of the Cori et al. and Wallinga and Teunis estimators (their equations 4.1 and 4.2) can be derived from the renewal equation model.

Originally developed in the context of population biology, the renewal equation is usually expressed as 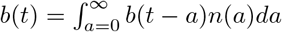, where *b*(*t*) is the number of births at time t and *n*(*a*) is fecundity at age *a*, scaled by the probability of surviving to age *a*. When used to describe epidemic dynamics, the renewal equation model is expressed in terms of *I*(*t*), the number of infections incident at time *t; S*(*t*), the susceptible fraction; and *w*(·), the generation interval distribution [29, 31, 32]. Note that *R_0_S*(*t*) = *R_t_*.

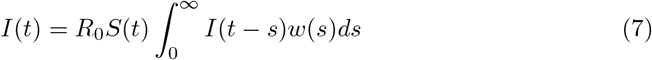

The difficulty is that the “intrinsic” generation interval of the renewal equation, which is the interval needed for accurate *R_t_* estimation, is conceptually and quantitatively different from the generation intervals observed in practice [31–33]. In the renewal equation, the generation interval describes the time distribution of all infectious contact events, whether or not the contacted individual is susceptible. (Note that a different factor in the equation, *S*(*t*), scales the probability of contacting a susceptible individual and causing a new infection.) The demographic analogy is that *w*(·) describes changes over time in the fecundity (infectiousness) of an index case, while *S*(*t*) determines whether offspring of that index case are viable. In practice, we only observe generation intervals between viable pairs. Thus, unlike the intrinsic generation interval, observed intervals are sensitive to changes in the rate of susceptible depletion and can be biased estimators of the intrinsic interval, and of *R_t_* [31–33]. A related challenge is that interventions such as contact tracing and self-isolation can limit transmission late in the course of infectiousness, shortening observed generation and serial intervals [32, 34, 35]. Methods for accurate estimation of the generation interval from contact tracing data involve adjusting for right truncation, and accounting for population susceptibility at the times transmission pairs are observed [32, 33].

The serial interval, defined as the time between symptom onset in an infector-infectee pair, is more easily observed than the generation interval and often used in its place. Although the serial and generation interval are often conflated, failure to understand the differences between these related quantities can bias *R_t_* estimates [21, 32]. The serial interval and the generation interval have the same mean, but usually different variance [36, 37], and the serial interval can be negative (e.g., for COVID-19 [38–40]), whereas the generation interval cannot [21, 32]. The intrinsic generation interval can be estimated from contact tracing data (i.e., estimates of the serial interval) [32, 33].

Fig. 3A illustrates the consequences of misspecifying the mean generation interval in the method of Cori et al. If the mean generation interval is set too high, *R_t_* values will typically be further from 1 than the true value—too high when *R_t_* > 1 and too low when *R_t_* < 1. If the mean is set too low, *R_t_* values will typically be closer to 1 than the true value. These biases are relatively small when *R_t_* is near the critical threshold of one but increase as *R_t_* takes substantially higher or lower values (Fig. 3). Therefore, biases from misspecification of the generation interval may be greatest early in an epidemic, when the sensitivity of high *R_t_* values to bias may be compounded by limited data and highly uncertain generation interval estimates.

**Fig 3.**
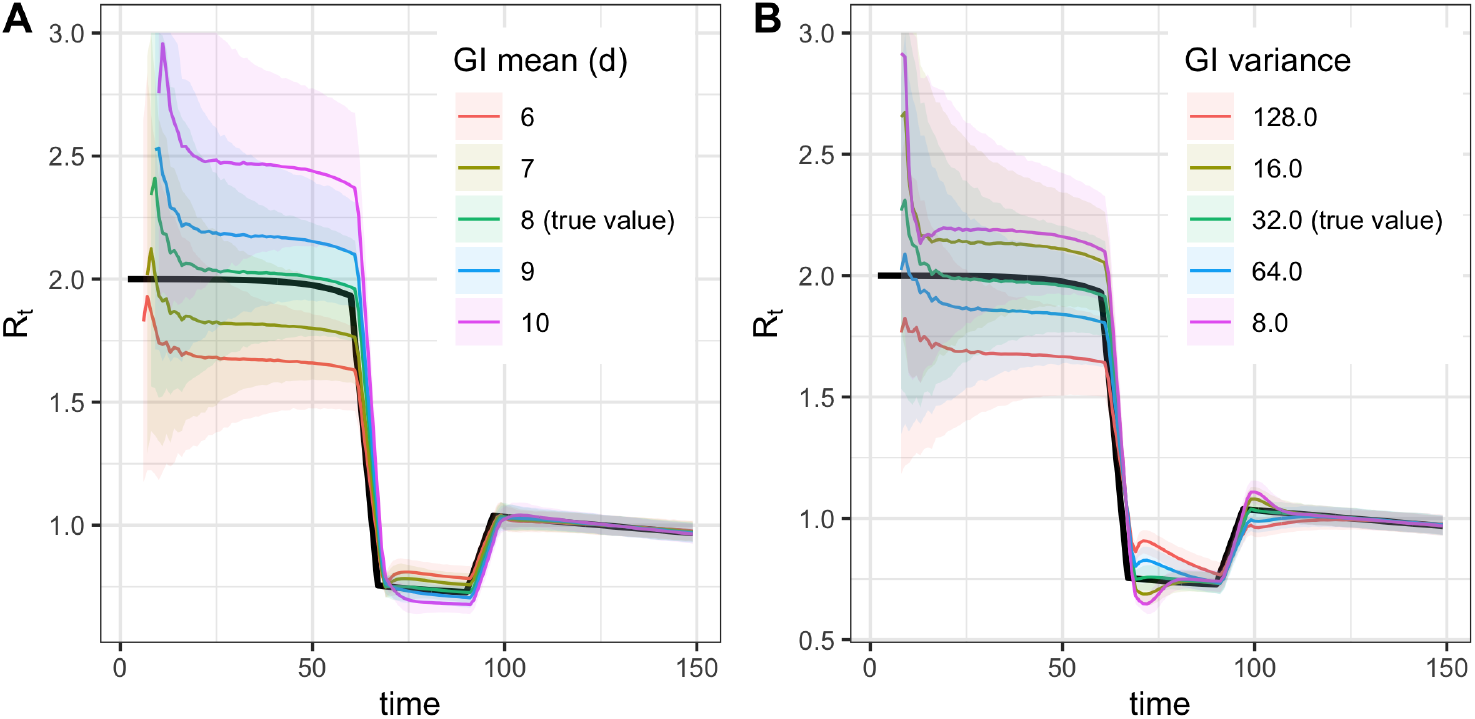
Biases from misspecification of the generation interval mean (A) or variance (B) Demonstrated using the method of Cori et al.

The consequences of misspecifying the form and variance of the generation interval distribution are illustrated in Fig. 2A and Fig. 3B. As explained above, biases in the Bettencourt and Ribeiro estimator arise entirely from misspecification of the form of the generation interval, even if the mean is correctly specified. Similarly, the accuracy of the Cori et al. and Wallinga and Teunis estimators in Fig. 2 B, C are contingent on perfect specification of the generation interval; in practice, if the mean, variance or form of a pathogen’s true generation interval is uncertain, *R_t_* estimates obtained using these methods can be biased.

EpiEstim [20] allows users to account for uncertainty in the mean and standard deviation of the generation interval by resampling over a range of plausible values [14, 20]. Similarly, Bayesian methods such as EpiNow2 [41] and the rt.live adaptation [6] of the Bettencourt and Ribeiro method allow users to specify the prior variance of the mean and standard deviation. Uncertainty around an incorrect value can widen the resulting 95% interval but will not shift the assumed central value toward the truth, and will not correct bias in the central *R_t_* estimates.

Joint estimation of both *R_t_* and the serial interval is possible, depending on data quality and magnitude of *R_t_* [12, 42, 43], and the EpiEstim [15, 20] package provides an off-the-shelf option for joint estimation. However, these off-the-shelf methods should be used with caution, as they estimate the observed serial interval, not the intrinsic generation interval, and do not account for changes over time in behavior or susceptibility.

### Summary

- The intrinsic generation interval is required to correctly define the relationship between *R_t_* and incident infections.
- The intrinsic generation interval is rarely observable, and care must be taken to estimate it from proxies such as the serial interval.

## Adjusting for delays

Estimating *R_t_* requires data on the daily number of new infections (i.e., transmission events). Due to lags in the development of detectable viral loads, symptom onset, seeking care, and reporting, these numbers are not readily available. All observations reflect transmission events from some time in the past. In other words, if d is the delay from infection to observation, then observations at time t inform *R_t−d_*, not *R_t_* (Fig. 4). Obtaining temporally accurate *R_t_* estimates thus requires assumptions about lags from infection to observation. If the distribution of delays can be estimated, then *R_t_* can be estimated in two steps: first by inferring the incidence time series from observations and then by inputting the inferred time series into an *R_t_* estimation method. Alternatively, the unobserved time series could be inferred simultaneously with *R_t_* or treated as a latent state. Such methods are now under development and available in a development version of the R package EpiNow2 [41, 44].

**Fig 4.**
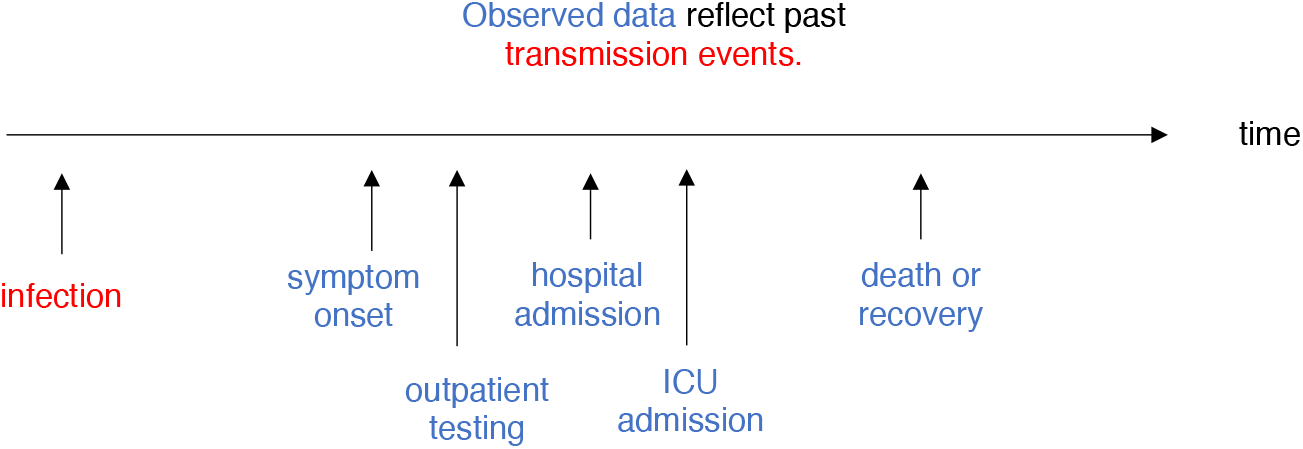
*R_t_* is a measure of transmission at time t. Observations after time t must be adjusted.

Two simple but mathematically incorrect methods for inference of unobserved times of infection have been applied to COVID-19: convolution and temporal shifts. The errors introduced by these methods may be tolerable if delays to observation are relatively short and not highly variable, and if *R_t_* is not rapidly changing. But when dealing with longer or more variable observation delays, or when aiming to infer the timing of changes in *R_t_* accurately, these methods may not be sufficient.

One method infers each individual’s time of infection by subtracting a sample from the delay distribution from each observation time. This is mathematically equivalent to convolving the observation time series with the reversed (backward) delay distribution, but convolution does not accurately infer the underlying time series of infections from observations [45-47]. The forward delay distribution has the effect of spreading out infections incident on a particular day across many days of observation. This blurring into the future is biologically realistic and reflects individual variation in disease progression and care seeking. To recover the original time series of infections from observations requires deblurring. Instead, as illustrated in (S1 Fig), backward convolution unrealistically spreads them out further. An unintended consequence of added blurring from backward convolution is that it can help smooth over weekend effects and other observation noise. But a crucial pitfall is that this blurring also smooths over true variation in *R_t_*: peaks, valleys, and changes in slope of the latent time series of infection events. Convolution and other approaches that blur or oversmooth can therefore prevent or delay detection of changes in *R_t_* and can impede accurate inference of the timing of these changes (Fig. 5C).

**Fig 5.**
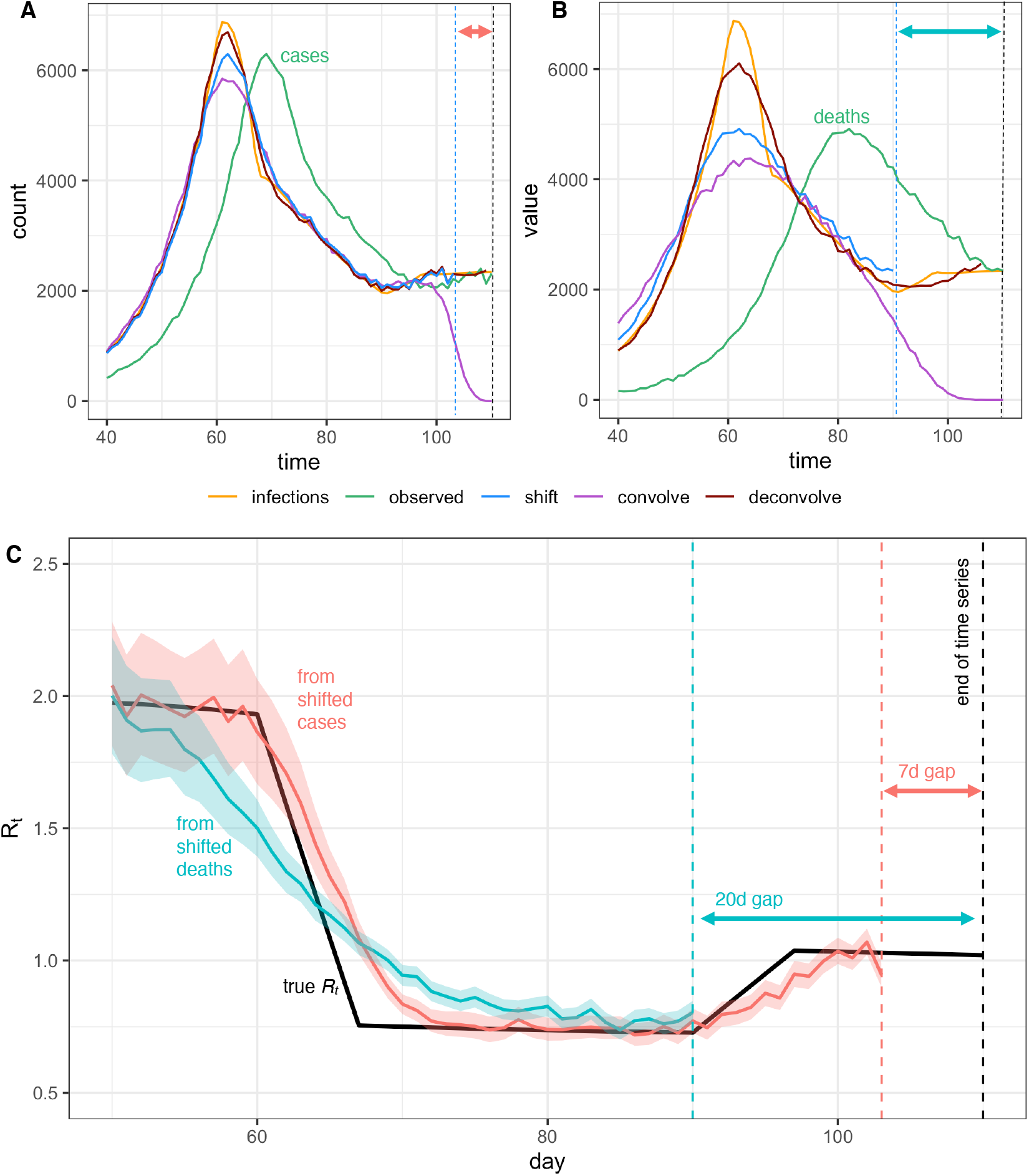
Pitfalls of simple methods to adjust for delays to observation when estimating *R_t_*. Infections back calculated from (A) observed cases or (B) observed deaths either by shifting the observed curve back in time by the mean observation delay (shift), by subtracting a random sample from the delay distribution from each individual time of observation (convolve), or by deconvolution (deconvolve), without adjustment for right truncation. Neither back-calculation strategy accurately recovers peaks or valleys in the true infection curve. The inferred infection curve is less accurate when the variance of the delay distribution is greater (B vs. A). (C) Posterior mean and credible interval of *R_t_* estimates from the Cori et al. method. Inaccuracies in the inferred incidence curves affect *R_t_* estimates, especially when *R_t_* is changing (here *R_t_* was estimated using shifted values from A and B). Finally we note that shifting the observed curves back in time without adjustment for right truncation leads to a gap between the last date in the inferred time series of infection and the last date in the observed data, as shown by the dashed lines and horizontal arrows in A-C.

The second simple-but-incorrect method to adjust for lags is to shift either the raw inputs (the observed time series) or outputs of *R_t_* estimation on the time axis. *R_t_* estimates obtained by applying the methods of Cori et al. or Bettencourt and Ribeiro to unadjusted data will lag the true instantaneous *R_t_* by roughly the mean delay from infection to observation. Because the case reproductive number leads the instantaneous reproductive number by roughly one generation interval, the unadjusted estimates obtained from the Wallinga and Teunis method will lag the true instantaneous reproductive number by roughly the mean delay to observation minus the mean generation interval. Unlike backward convolution, temporal shifting does not further blur the observed time series. Thus, if the mean delay is known accurately, this method is preferable to subtracting samples from the delay distribution (Fig. 5 A, B). However, shifting the input time series does not undo the blurring effect of the original delay, which, like backward convolution can impede accurate inference of changes in *R_t_*. Shifting inputs or *R_t_* estimates by a fixed amount also fails to account for realistic uncertainty in the true mean delay, which will not be known exactly and might change over time.

More reliable methods to reconstruct the incidence time series are now under development. Given a known delay distribution, one potential solution is to infer the unlagged signal using maximum-likelihood deconvolution. This method was applied to AIDS cases, which feature long delays from infection to observation [47], and in the reconstruction of incidence from mortality times series for the 2009 H1N1 pandemic [45]. It is now being applied to COVID-19 [8, 48]. Fig. 5 shows an example of deconvolution applied following the methods in [45]. The method of [47] is implemented in the backprojNP function within the surveillance R package [49, 50]. In principle, deconvolution can more accurately estimate the latent time series than temporal shifting or backward convolution, but the method is sensitive to misspecification of the mean, variance or form of the delay distribution, and the stringency of the stopping condition of the deconvolution algorithm. It can also be difficult to quantify uncertainty in the deconvolved time series [41] and to implement deconvolution while adjusting for right truncation.

A potential alternative to deconvolution is *R_t_* estimation models that include forward delays to observation in the inference process or that treat the time series of infections as latent states. Such methods are in development within the R package EpiNow2 [41]. An additional advantage of inferring the time series of infections jointly with *R_t_* is seamless integration of various sources of uncertainty, e.g., in *R_t_* and reporting. By comparison, the two-step approach of first transforming the observed time series and then calculating *R_t_* requires users to propagate uncertainty from the back-calculation step into the *R_t_* estimation step. A final advantage of latent state methods is that they could in theory facilitate inference from multiple data streams simultaneously. For example, by assuming that cases, hospitalizations, and deaths all arise from a common infection process, these methods might be able to infer the incident time series of infections more accurately and precisely, potentially while also estimating delays and changes in ascertainment for specific data sources (e.g., outpatient cases).

Deconvolution or *R_t_* estimation methods that include a forward observation process are particularly useful when delays to observation are relatively long and variable, and in analyses that require accurate inference of the timing and speed of changes in *R_t_*. If delays to observation are relatively short, or if *R_t_* is not substantially changing, then deconvolution may not be necessary. For example, when working with synthetic case data in which the mean delays to observation are short and known accurately, the underlying infection curve (Fig. 5A) and underlying *R_t_* values (Fig. 5C) can be recovered with reasonable accuracy simply by shifting the observed time series. But longer and more variable delays to observation worsen inference of the underlying incidence curve (Fig. 5B). In turn, this makes it more difficult to infer the speed and timing of abrupt changes in *R_t_* and to relate those changes to policies, behaviors or other extrinsic epidemic drivers at specific points in time. For example, simply shifting a times series of observed deaths by the mean delay does not accurately recover the underlying curves of infections or *R_t_* (Fig. 5B, C). The marginal value of deconvolution or other methods that infer the unobserved time series of infections is greater when delays to observation are longer and more variable, and when *R_t_* is changing.

Another advantage of working with observations nearer the time of infection, such as times of symptom onset among newly symptomatic individuals, is that they provide more information about recent transmission events and therefore allow *R_t_* to be estimated in closer to real-time (Fig. 5C) [46]. Of course, this advantage could be offset by sampling biases and reporting delays. Users will need to balance data quality with the length of the observation delay when selecting inputs.

Further investigation is needed to determine the best methods for inferring infections from observations if the underlying delay distribution is uncertain. If the delay distribution is severely misspecified, all three approaches (deconvolution, shifting by the mean delay, or convolution) will incorrectly infer the timing of changes in incidence. Inthis case, methods such as deconvolution or shifting by the mean delay might more accurately estimate the magnitude of changes in *R_t_* but at the cost of spurious precision in the inferred timing of those changes. Ideally, the delay distribution could be inferred jointly with the underlying times of infection or estimated as the sum of the incubation period distribution and the distribution of delays from symptom onset to observation (e.g. from line-list data).

### Summary

- Estimating the instantaneous reproductive number requires data on the number of new infections (i.e., transmission events) over time. These inputs must be inferred from observations using assumptions about delays between infection and observation.
- Inferring the unlagged time series of infections using deconvolution, or within an *R_t_* estimation model that includes forward delays, can improve accuracy.
- A less accurate but simpler approach is to shift the observed time series by the mean delay to observation. If the delay to observation is not highly variable, and if the mean delay is known exactly, the error of this approach may be tolerable. A key disadvantage is that shifting by a fixed amount of time does not account for uncertainty or individual variation in delay times.
- Sampling from the delay distribution to impute individual times of infection from times of observation accounts for uncertainty but blurs peaks and valleys in the underlying incidence curve, which in turn compromises the ability to rapidly detect changes in *R_t_*.

## Adjusting for right truncation

Near real-time estimation requires not only inferring times of infection from the observed data but also adjusting for missing observations of recent infections. The absence of recent infections is known as “right truncation”. Without adjustment for right truncation, the number of recent infections will appear artificially low because they have not yet been reported [4, 30, 51–55]. Thus, adjusting for right truncation is particularly important in analyses with the goal of near real-time *R_t_* estimation.

Figure 5 illustrates the consequences of failure to adjust for right truncation when inferring times of infection from observations. Subtracting the mean observation delay m from times of observation (“shift” method in Fig. 5A, B) leaves a gap of m days between the last date in the inferred infection time series and the last date in the observed data. This hampers recent *R_t_* estimation (Fig. 5C). Inferring the underlying times of infection by subtracting samples from the delay distribution (“convolve” method in Fig. 5A, B) dramatically underestimates the number of infections occurring in the last few days of the time series.

The simplest approach is to drop estimates on the last few dates, or to flag them as unreliable [8]. But many methods are available to adjust for right truncation, which can improve real-time analysis [41, 51-56]. These methods infer based on past reporting delays the total number of infections, observed and not-yet-observed, at the end of the time series.

In short, accurate near real-time *R_t_* estimation requires both inferring the infection time series from recent observations and adjusting for right truncation. Errors in either step could amplify errors in the other. Joint inference approaches for near real-time *R_t_* estimation, which simultaneously infer times of infection and adjust for right truncation, are now in development [41].

### Summary

- At the end of a truncated time series, some infections will not yet have been observed. Infer the missing data to obtain accurate recent *R_t_* estimates.

## Accounting for incomplete observation

The effect of incomplete case observation on *R_t_* estimation depends on the observation process. If the fraction of infections observed is constant over time, *R_t_* point estimates will remain accurate and unbiased despite incomplete observation [12, 14, 43, 57, 58]. Data obtained from carefully designed surveillance programs might meet these criteria. But even in this best-case scenario, because the estimation methods reviewed here assume all infections are observed, confidence or credible intervals obtained using these methods will not include uncertainty from incomplete observation. Without these statistical adjustments, practitioners and policy makers should beware false precision in reported *R_t_* estimates.

Sampling biases will also bias *R_t_* estimates [57]. COVID-19 test availability, testing criteria, interest in testing, and even the fraction of deaths reported [59] have all changed over time. If these biases are well understood, it might be possible to adjust for them when estimating *R_t_*. Observed hospitalizations and deaths may be less sensitive to changes in test availability and testing effort, but may be more sensitive to other factors, such as hospital bed availability. Hospitalizations and deaths will also vary in their representativeness of mean transmission rates, depending on which age groups are being infected. No matter what data is used, one potential solution is to flag *R_t_* estimates as potentially biased in the few weeks following known changes in data collection or reporting. At a minimum, practitioners and policy makers should understand how the data underlying *R_t_* estimates were generated and whether they were collected under a standardized testing protocol.

### Summary

- *R_t_* point estimates will remain accurate given imperfect observation of cases if the fraction of cases observed is time-independent and representative of a defined population. But even in this best-case scenario, confidence or credible intervals will not accurately measure uncertainty from imperfect observation.
- Changes over time in the type or fraction of infections observed can bias *R_t_* estimates. Structured surveillance with fixed testing protocols can reduce or eliminate this problem.

## Smoothing windows

Because *R_t_* estimators incorrectly assume all infections are observed, day-of-week reporting effects and stochasticity in the number of observations per day can cause spurious variability in *R_t_* estimates, especially if the number of observations per day is low [14]. The Cori method incorporates a sliding window to smooth *R_t_* estimates, but the size of the smoothing window can affect both the temporal and quantitative accuracy of estimates. Larger windows effectively increase sample size by drawing information from multiple time points but blur what may be biologically meaningful changes in *R_t_*. Some smoothing approaches can also cause *R_t_* estimates to lead or lag the true value (Fig. 6).

**Fig 6.**
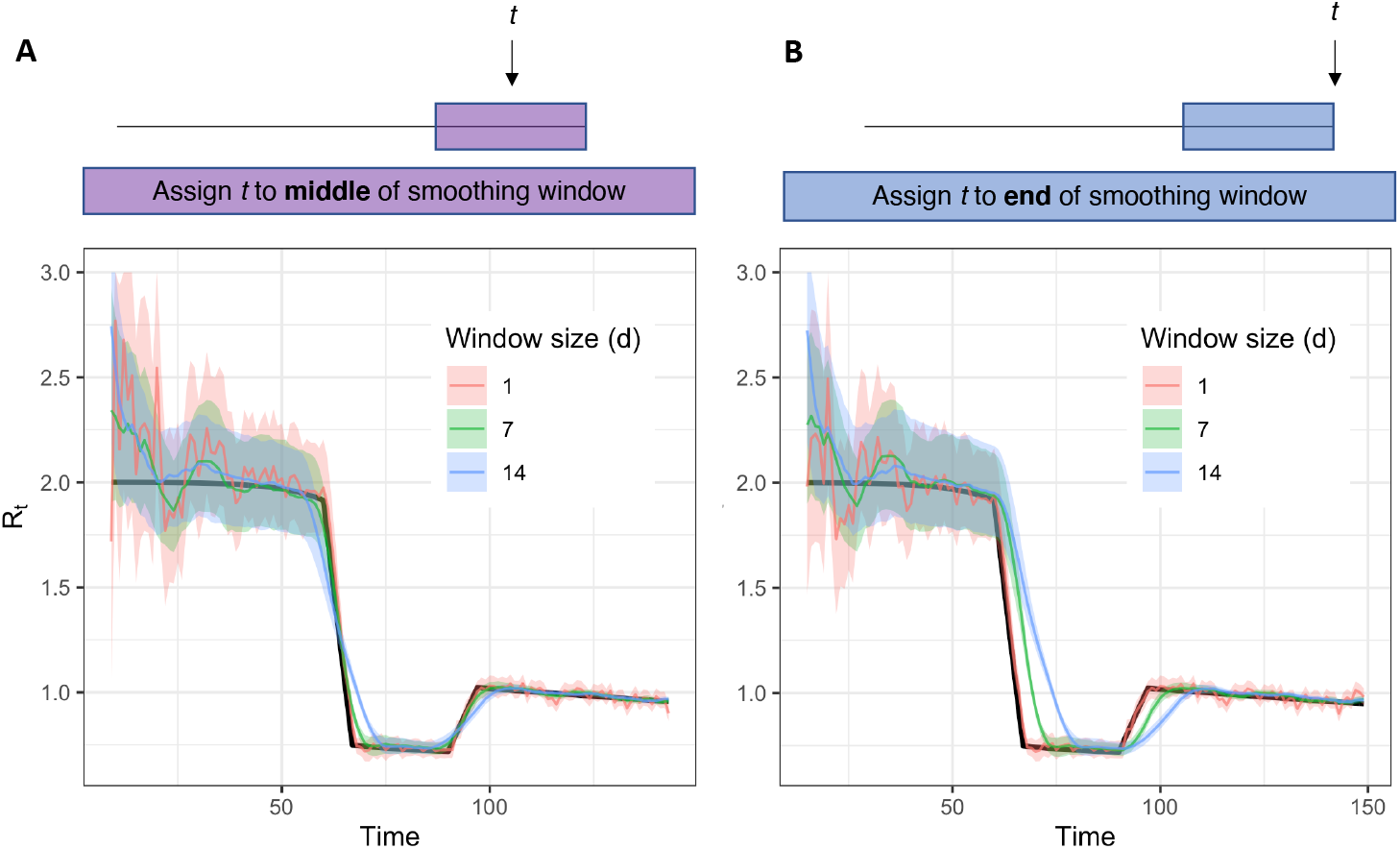
Accuracy of *R_t_* estimates given smoothing window width and location of *t* within the smoothing window. Estimates were obtained using synthetic data drawn from the *S* → *E* transition of a stochastic SEIR model (inset) as an input to the method of Cori et al. Colored estimates show the posterior mean and 95% credible interval. Black line shows the exact instantaneous *R_t_* calculated from synthetic data.

Lags can be particularly severe when using the conventions suggested by Cori et al., in which *R_t_* is reported on the last date in a given window, rather than on the middle date. Although this convention returns *R_t_* estimates to the last date in the time series, which is convenient for real-time estimation, *R_t_* estimates reported at the end of a window are based entirely on data from the past and therefore lag the instantaneous *R_t_* (Fig 6B). Instead, we recommend reporting *R_t_* at the midpoint of the smoothing window (Fig 6A), which produces estimates that are more accurately oriented in time. An apparent disadvantage is that a centered window precludes *R_t_* estimation on the last *w/2* time units, where *w* is the width of the window. However, we argue that temporal accuracy is preferable, and that if daily counts are low, failure to produce estimates on the last few days in the time series realistically acknowledges uncertainty about recent trends. Thus, for SARS-CoV-2 and other pathogens with short timescales of infection, near real-time *R_t_* estimation requires large enough daily counts to permit a small window (e.g., a few days).

Although the sliding window increases statistical power to infer *R_tl_* it does not by itself accurately calculate confidence intervals. Thus, underfitting and overfitting are possible. The risk of overfitting in the Cori method is determined by the length of the time window that is chosen. In other words, there is a trade-off in window length between picking up noise with very short windows and over-smoothing with very long ones. To avoid this, one can choose the window size based on short-term predictive accuracy, for example using leave-future-out validation to minimize the one-step-ahead log score [60]. Proper scoring rules such as the Ranked Probability Score can be used in the same way, and a time-varying window size can be chosen adaptively [41].

### Summary

- If *R_t_* appears to vary abruptly due to underreporting, a wide smoothing window can help resolve *R_t_*. However, wider windows can also lead to lagged or inaccurate *R_t_* estimates.
- If a wide smoothing window is needed, consider reporting *R_t_* for t corresponding to the middle of the window.
- To avoid overfitting, choose a smoothing window based on short-term predictive accuracy [60] or use an adaptive window [41].

## Conclusion

We tested the accuracy of several methods for *R_t_* estimation in near real-time and recommend the methods of Cori et al. [14], which are currently implemented in the R package EpiEstim [20]. The Cori et al. method estimates the instantaneous rather than the case reproductive number and is conceptually appropriate for near real-time estimation. The method uses minimal parametric assumptions about the underlying epidemic process and can accurately estimate abrupt changes in the instantaneous reproductive number using ideal, synthetic data.

Most epidemiological data are not ideal, and statistical adjustments are needed to obtain accurate and timely *R_t_* estimates. First, to obtain timely and temporally accurate *R_t_* estimates, considerable pre-processing is needed to infer the underlying time series of infections (i.e., transmission events) from delayed observations and to adjust for right truncation. Best practices for this inference are still under investigation, especially if the delay distribution is uncertain. The smoothing window must also be chosen carefully, potentially adaptively, and daily counts must be sufficiently high for changes in *R_t_* to be resolved on short timescales. To avoid biases in *R_t_* estimates, the generation interval distribution must be estimated and specified accurately. Finally, to avoid false precision in *R_t_*, uncertainty arising from delays to observation, from adjustment for right truncation, and from imperfect observation must be propagated. The functions provided in the EpiEstim package quantify uncertainty arising from the *R_t_* estimation model but currently not from uncertainty arising from imperfect observation or delays.

Work is ongoing to determine how best to infer infections from observations and to account for all relevant forms of uncertainty when estimating *R_t_*. Some useful extensions of the methods provided in EpiEstim have already been implemented in the R package EpiNow2 [41, 44], and further updates to this package are planned as new best practices become established.

But even the most powerful inferential methods, extant and proposed, will fail to estimate *R_t_* accurately if changes in sampling are not known and accounted for. If testing shifts from more to less infected subpopulations, or if test availability shifts over time, the resulting changes in case numbers will be ascribed to changes in *R_t_*. Thus, structured surveillance also belongs at the foundation of accurate *R_t_* estimation. This is an urgent problem for near real-time estimation of *R_t_* for COVID-19, as case counts in many regions derive from clinical testing outside any formal surveillance program. Deaths, which are more reliably sampled, are lagged by 2-3 weeks and still subject to biases in underreporting. The establishment of sentinel populations (e.g., outpatient visits with recent symptom onset) for *R_t_* estimation could thus help rapidly identify the effectiveness of different interventions and recent trends in transmission.

## Data Availability

This is a theoretical study analyzing only synthetic data. All code to generate and analyze the data is publicly available on Github (url provided within the text).

https://github.com/cobeylab/Rt_estimation

## Code availability

All code for analysis and figure generation is available at https://github.com/cobeylab/Rt_estimation.

## Supporting information

**S1 Fig.**
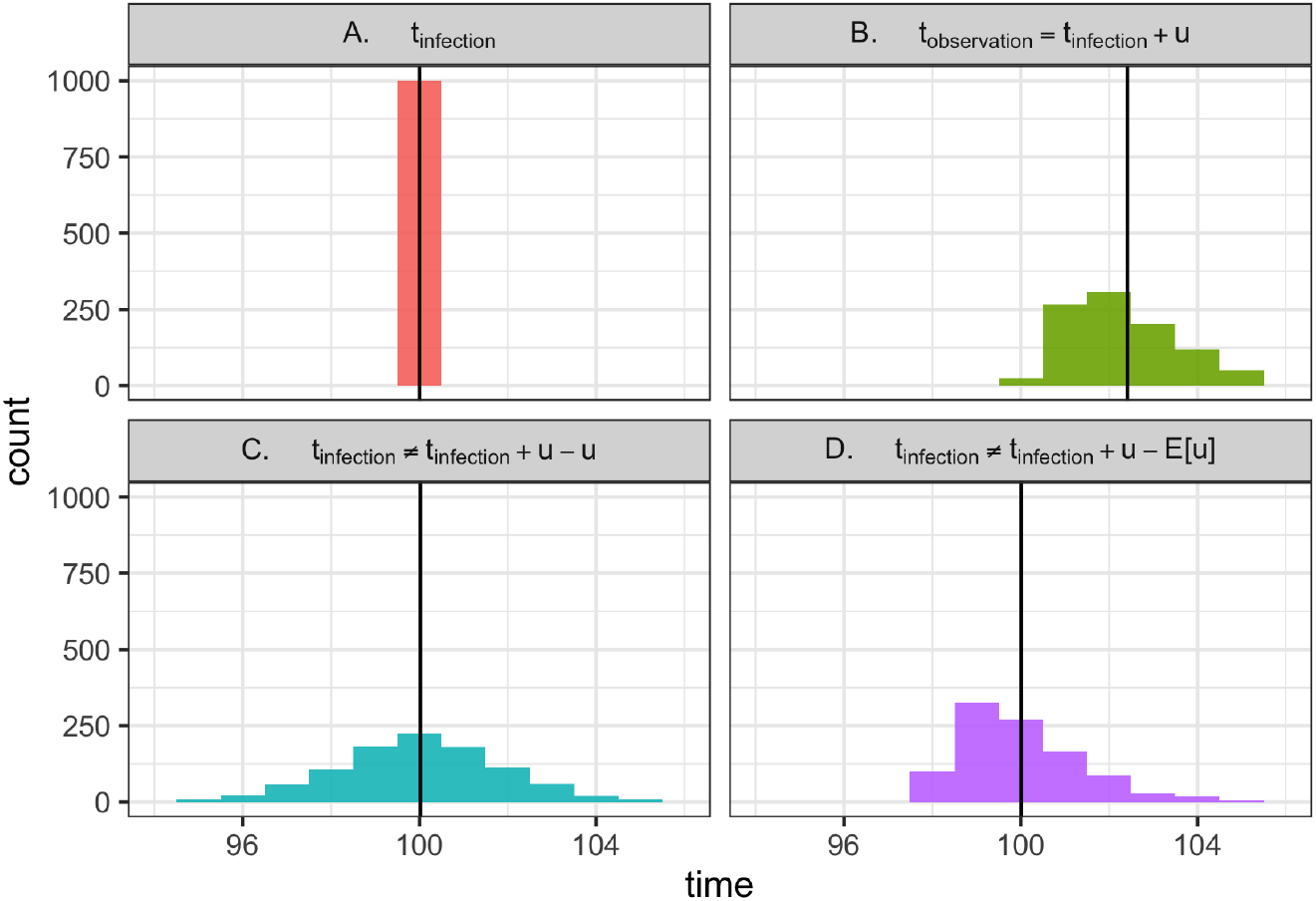
Why is deconvolution needed to recover latent times of infection? (A) Consider 1000 individuals, all infected at time 100. (Vertical line shows the mean). (B) Now consider the times at which these individuals are observed. Logically, *t*_observation_ = *t*_infected_ + *u*, where *u* is a random variable describing the delay between infection and observation. Mathematically, this is a convolution of the infection time and the delay distribution. Because u has non-zero variance, observation times are not only shifted into the future but also are blurred across many dates. This blurring is biologically realistic; due to variability in disease progression and care seeking, individuals with the same date of infection will not necessarily be observed at the same time. (C) Using the observations in B, we aim to recover the latent times of infection shown in A. Doing so would require not only shifting into the past but also removing the variance introduced by the observation process, which can be achieved by deconvolution. Instead, as demonstrated here, a common strategy is to subtract *u* from the times of observation, effectively repeating the convolution shown in B, but this time moving backward in time rather than forward. This is not the correct inverse operation. It fails to remove variance introduced by the observation process (the forward convolution) and adds new, biologically unrealistic variance, further blurring the inferred times of infection. (D) Shifting the times of observation by the mean delay *E*[*u*] is also incorrect, as it does not remove the variance from the forward convolution in B. But if the mean delay time is known exactly, this approach is preferable to C, as it avoids adding even more variance. Ultimately, deconvolution methods would be needed to recover A from the observations in B while properly accounting for uncertainty.

**S2 Fig.**
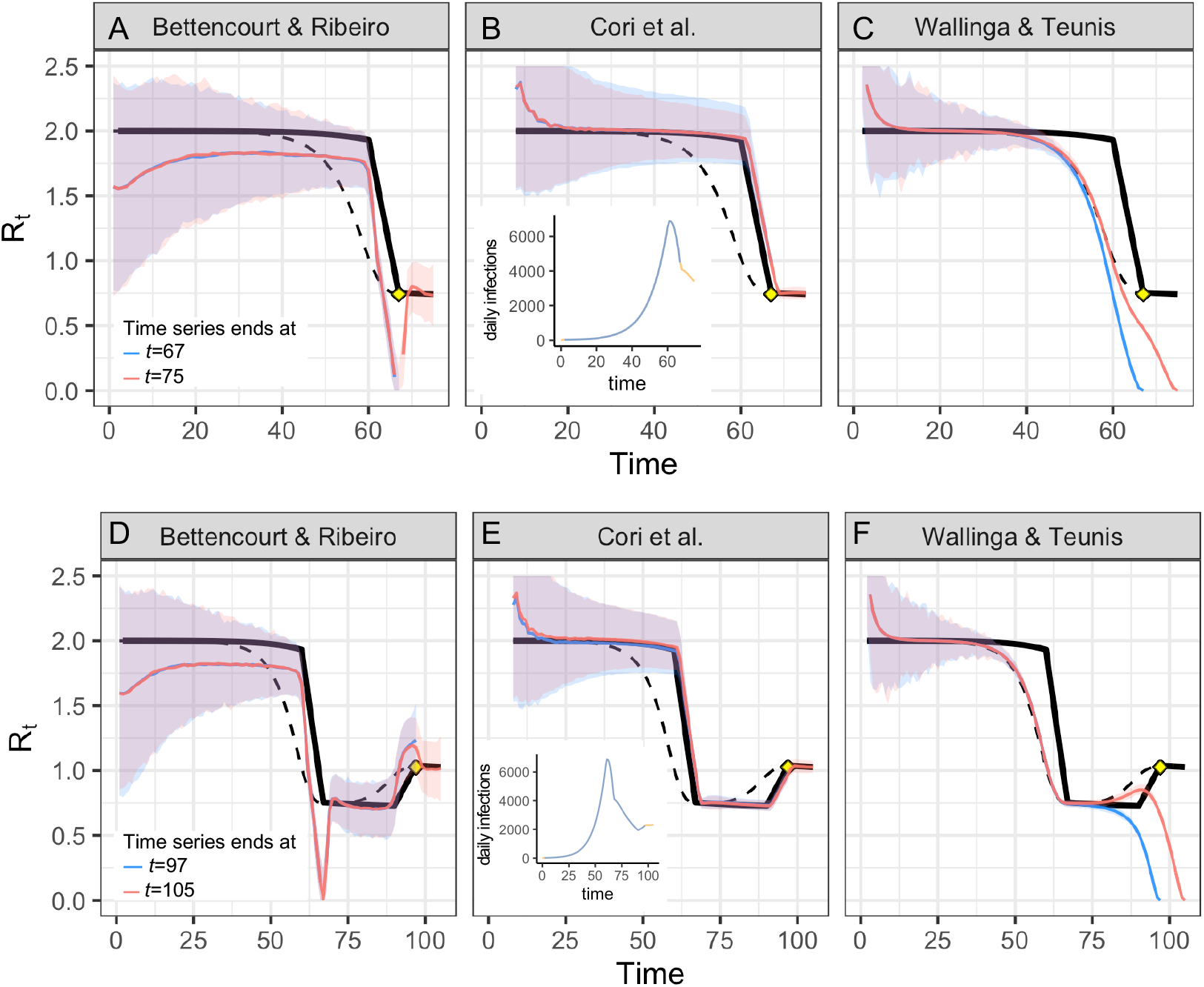
real-time accuracy when *R_t_* is rising or falling. (A-C) Alternate version of Fig. 2 in which the time series ends on the day *R_t_* first hits its minimum value after falling abruptly (time 67, yellow point), or eight days after the changepoint (time 75). (D-F) The time series ends on the day *R_t_* stops rising (time 97, yellow point), or eight days later (time 105). Estimates of the instantaneous reproductive number (A, B, D, E) remain accurate to the end of the time series, and estimates do not change as new observations become available in the 8 days following the changepoint. As in the main text, estimates of the unadjusted case reproductive number (C, F) depend on data from not-yet-observed time points. These estimates become more accurate as new observations are added to the end of the time series (orange vs. blue). Methods to infer the number of not-yet-observed infections can help make estimates of the case reproductive number more accurate in real-time [4, 30]. All panels show fits to the time series of new infections, and assume all infections are observed instantaneously. Solid black line shows the instantaneous reproductive number, and dashed black line shows the case reproductive number. Colored lines and confidence region show posterior mean and 95% credible interval (A, B, D, E) or maximum likelihood estimate and 95% confidence interval (C, F).

**S3 Fig.**
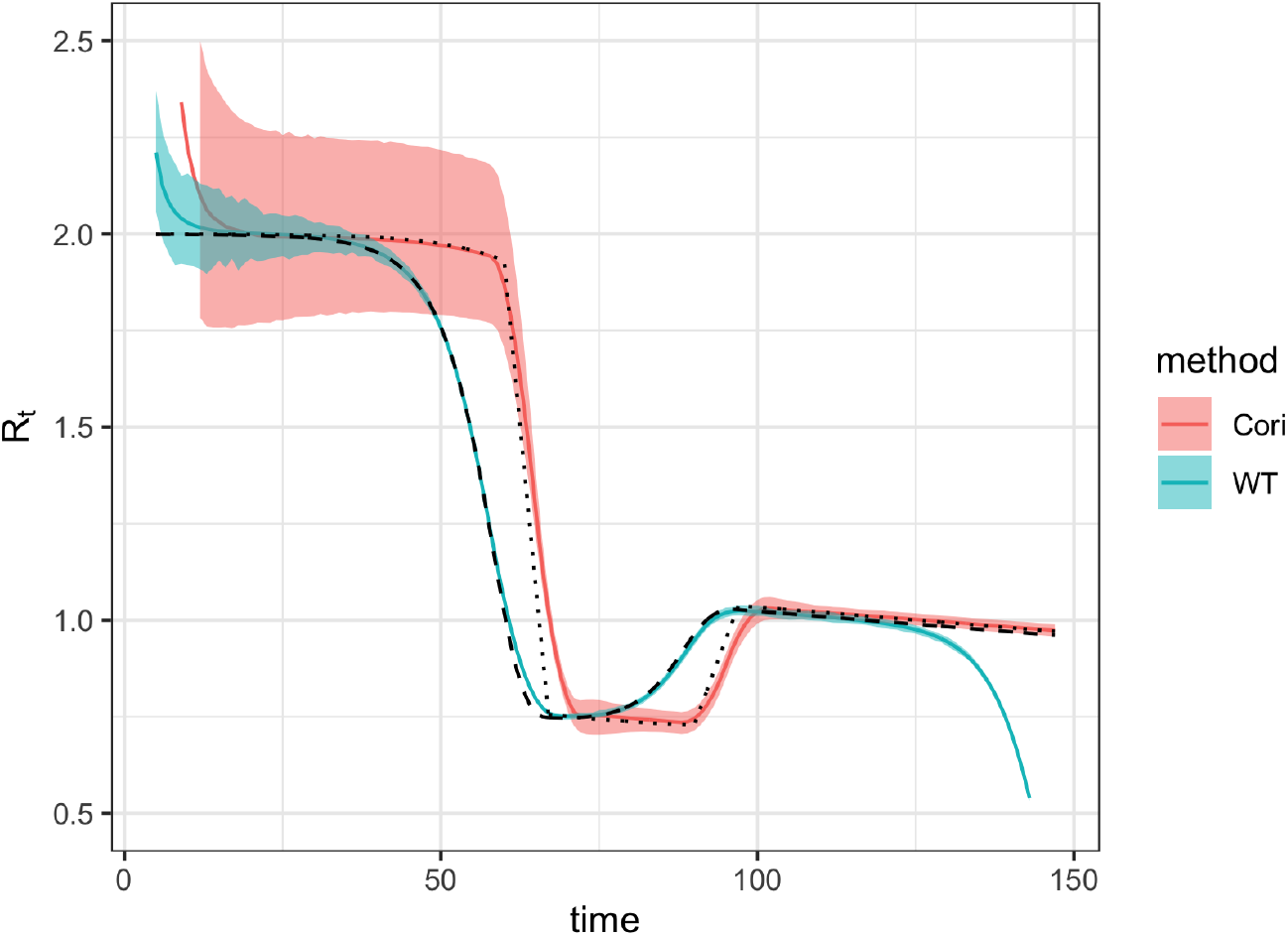
Smoothed estimates of Cori et al. and Wallinga and Teunis. Both were estimated using a 7-day smoothing window on a synthetic time series of new infections, observed without delay. The estimates of Cori et al. and Wallinga and Teunis are similar in shape when smoothed, but the estimate of Wallinga and Teunis (the case reproductive number) leads that of Cori et al. (the instantaneous reproductive number) by roughly 8 days, or the mean generation interval. Solid colored lines and confidence regions show the posterior mean and 95% credible interval (Cori et al.) or maximum likelihood estimate and 95% confidence interval (Wallinga and Teunis). Dotted and dashed lines show the exact instantaneous reproductive number and case reproductive number, respectively.

## Acknowledgements

We are grateful to Michael Höhle for helpful comments. KG was supported by the James S. McDonnell Foundation. LM was supported by the National Institute Of General Medical Sciences of the National Institutes of Health under Award Number F32GM134721. ML acknowledges support from the Morris-Singer Fund and from Models of Infectious Disease Agent Study (MIDAS) cooperative agreement U54GM088558 from the National Institute Of General Medical Sciences. The content is solely the responsibility of the authors and does not necessarily represent the official views of the National Institute Of General Medical Sciences or the National Institutes of Health. LFW acknowledges support from the National Institutes of Health (R01 GM122876). SA, JH, SM, JM, NIB, KS, RNT, SF acknowledge funding from the Wellcome Trust (210758/Z/18/Z). Thanks to Christ Church (Oxford) for funding via a Junior Research Fellowship (RNT). This project has been funded in whole or in part with Federal funds from the National Institute of Allergy and Infectious Diseases, National Institutes of Health, Department of Health and Human Services, under CEIRS Contract No. HHSN272201400005C (SC).

